# Explainable AI for Retinal Prostheses: Predicting Electrode Deactivation from Routine Clinical Measures

**DOI:** 10.1101/2021.03.07.21253092

**Authors:** Zuying Hu, Michael Beyeler

## Abstract

To provide appropriate levels of stimulation, retinal prostheses must be calibrated to an individual’s perceptual thresholds (‘system fitting’). Nonfunctional electrodes may then be deactivated to reduce power consumption and improve visual outcomes. However, thresholds vary drastically not just across electrodes but also over time, thus calling for a more flexible electrode deactivation strategy. Here we present an explainable artificial intelligence (XAI) model fit on a large longitudinal dataset that can 1) predict at which point in time the manufacturer chose to deactivate an electrode as a function of routine clinical measures (‘predictors’) and 2) reveal which of these predictors were most important. The model predicted electrode deactivation from clinical data with 60.8% accuracy. Performance increased to 75.3% with system fitting data, and to 84% when thresholds from follow-up examinations were available. The model further identified subject age and time since blindness onset as important predictors of electrode deactivation. An accurate XAI model of electrode deactivation that relies on routine clinical measures may benefit both the retinal implant and wider neuroprosthetics communities.

## I. INTRODUCTION

To provide appropriate levels of stimulation, retinal prostheses must be calibrated to each subject’s amount of electrical current needed to elicit visual responses (*perceptual threshold*). In the case of the Argus II Retinal Prosthesis System (Second Sight Medical Products, Inc.) [1], this process is part of *system fitting*, where perceptual thresholds are combined with electrode impedance measurements to populate subject-specific lookup tables that determine how the grayscale values of an image recorded by the external camera are translated into electrical stimuli. Nonfunctional electrodes are then deactivated, either because impedance measurements indicated an open or short circuit, or because no perceptual threshold below the charge density limit could be measured. Because this is a time-consuming process (each electrode requiring *∼* 100 trials of a behavioral detection task [2]), fitting sessions are thereafter limited to annual checks.

However, perceptual thresholds vary drastically not just across electrodes but also over time [2]–[5], thus calling for a more flexible electrode deactivation strategy. Thresholds often undergo sudden and large fluctuations that can last several weeks and cannot be explained by gradual changes in the implant-tissue interface [5]. Whereas the cochlear implant community has developed electrode deactivation strategies informed by recent device performance [6]–[8], no such strategy exists for retinal implant recipients. Repeated system fitting would be time-consuming at best and quickly become infeasible as new retinal prostheses are being developed that feature thousands of electrodes.

To address these challenges, we developed an explainable artificial intelligence (XAI) model that could 1) predict at which point in time an individual Argus II electrode was deactivated as a function of routinely collected clinical measures (‘predictors’), and 2) reveal which of these predictors were most important. Previous studies have focused on linear models [3], [4], which provide easily interpretable model parameters, but are often not powerful enough to fit the data. Machine learning (ML) models based on deep learning may offer state-of-the-art prediction accuracy, but are ‘black boxes’ whose predictions are inscrutable, and hence not actionable in a clinical setting. On the other hand, XAI relies on ML approaches that can explain why a certain prediction was made while maintaining high accuracy [9].

## II. METHODS

### A. Dataset

We analyzed a longitudinal dataset of 5, 496 perceptual thresholds and electrode impedances measured on 627 electrodes in 12 Argus II patients (Table I; for demographic information see [3], [10], [11]). The data was collected from 2007 *−* 2018 during 285 sessions conducted at 7 different implant centers located across the United States, the United Kingdom, France, and Switzerland. For all subjects, threshold measurements were available for a majority of the 60 electrodes in the array, measured 14 *−* 69 times over the lifetime of the device. Whereas only a handful electrodes were deactivated during system fitting (labeled ‘SF’ in Table I), most electrodes were at least temporarily deactivated over the life time of the device (labeled ‘LT’).

**TABLE I.** Prevalence of Argus II Electrode Deactivation

| Subjects | Data points | Sessions | Measured electrodes | Deactivated electrodes |  |
| --- | --- | --- | --- | --- | --- |
|  |  |  |  | SF | LT |
| 12-001 | 841 | 43 | 51 | 1 | 41 |
| 12-004 | 320 | 32 | 43 | 1 | 40 |
| 12-005 | 912 | 31 | 56 | 2 | 5 |
| 14-001 | 260 | 15 | 44 | 2 | 39 |
| 17-002 | 366 | 30 | 52 | 2 | 51 |
| 51-001 | 269 | 19 | 54 | 8 | 44 |
| 51-003 | 243 | 14 | 55 | 16 | 53 |
| 51-009 | 337 | 12 | 54 | 1 | 6 |
| 52-001 | 605 | 24 | 60 | 0 | 0 |
| 52-003 | 435 | 19 | 50 | 4 | 48 |
| 61-004 | 367 | 20 | 57 | 3 | 51 |
| 71-002 | 541 | 69 | 51 | 1 | 41 |
| **Total** | **5496** | **285** | **627** | **43** | **419** |

SF: system fitting, LT: life time
<sup>1</sup>Zuying Hu is with the Department of Computer Science, University of California, Santa Barbara, CA 93106, USA
<sup>2</sup>Michael Beyeler is with the Department of Computer Science and with the Department of Psychological & Brain Sciences, University of California, Santa Barbara, CA 93106, USA

A subset of the data was previously collected as part of the Argus II Feasibility Protocol (Clinical Trial ID: NCT00407602). Our study, which did not involve human subjects research, was exempt from IRB approval.

### B. Feature Engineering

To prepare the raw data for ML, we combined threshold and impedance values with clinical data crowd-sourced from the literature and performed feature engineering. The resulting feature correlation matrix is shown in Fig. 1, with each feature described in Table II and in more detail below.

**Fig. 1.**
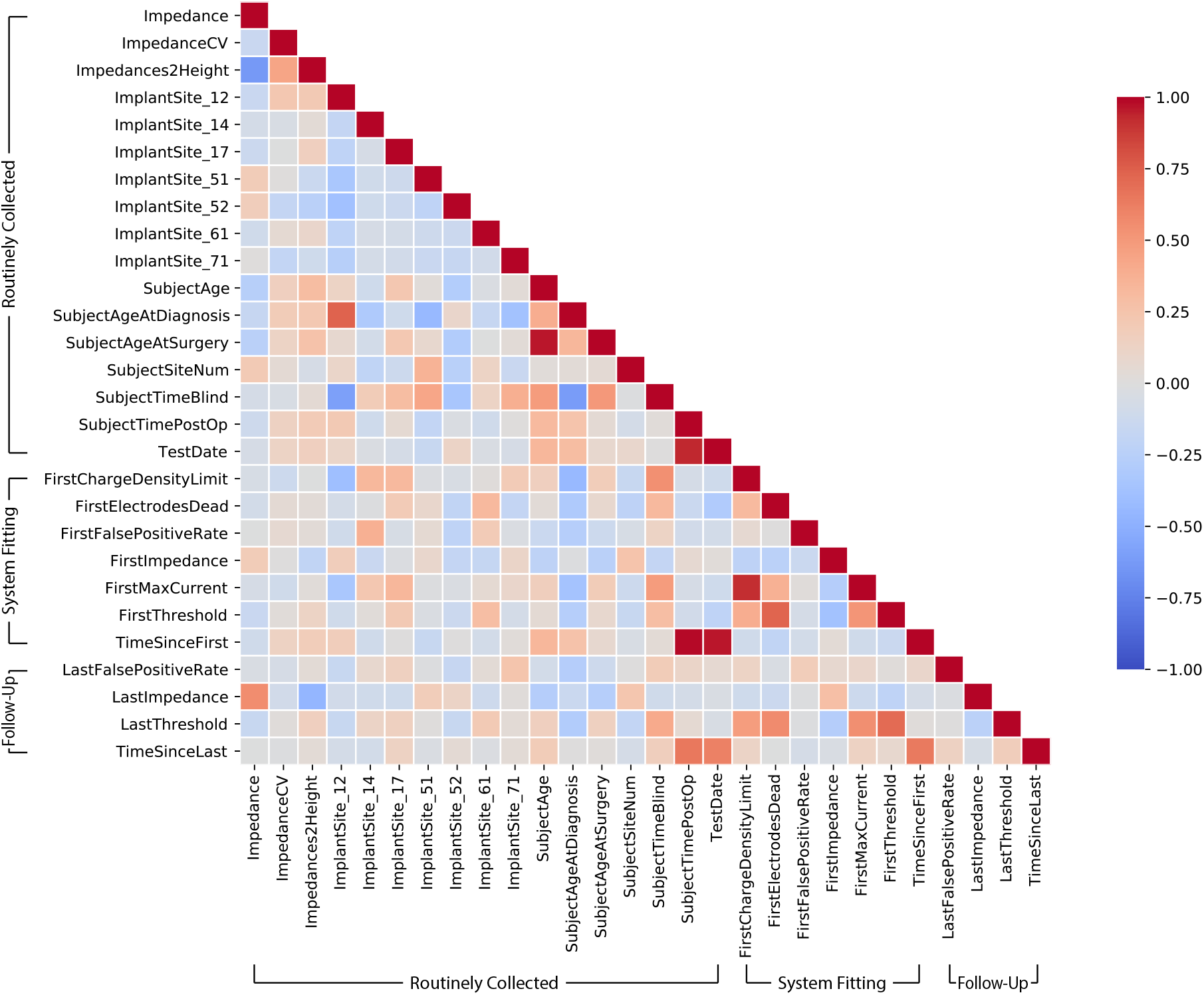
Feature correlation matrix: Heatmap of Pearson correlation coefficient for all feature pairs. Note that some timestamp-related features (e.g., ‘TestDate’, ‘SubjectTimePostOp’) appear highly correlated, which may impede the performance of linear models, but not necessarily tree–based models.

**TABLE II.**
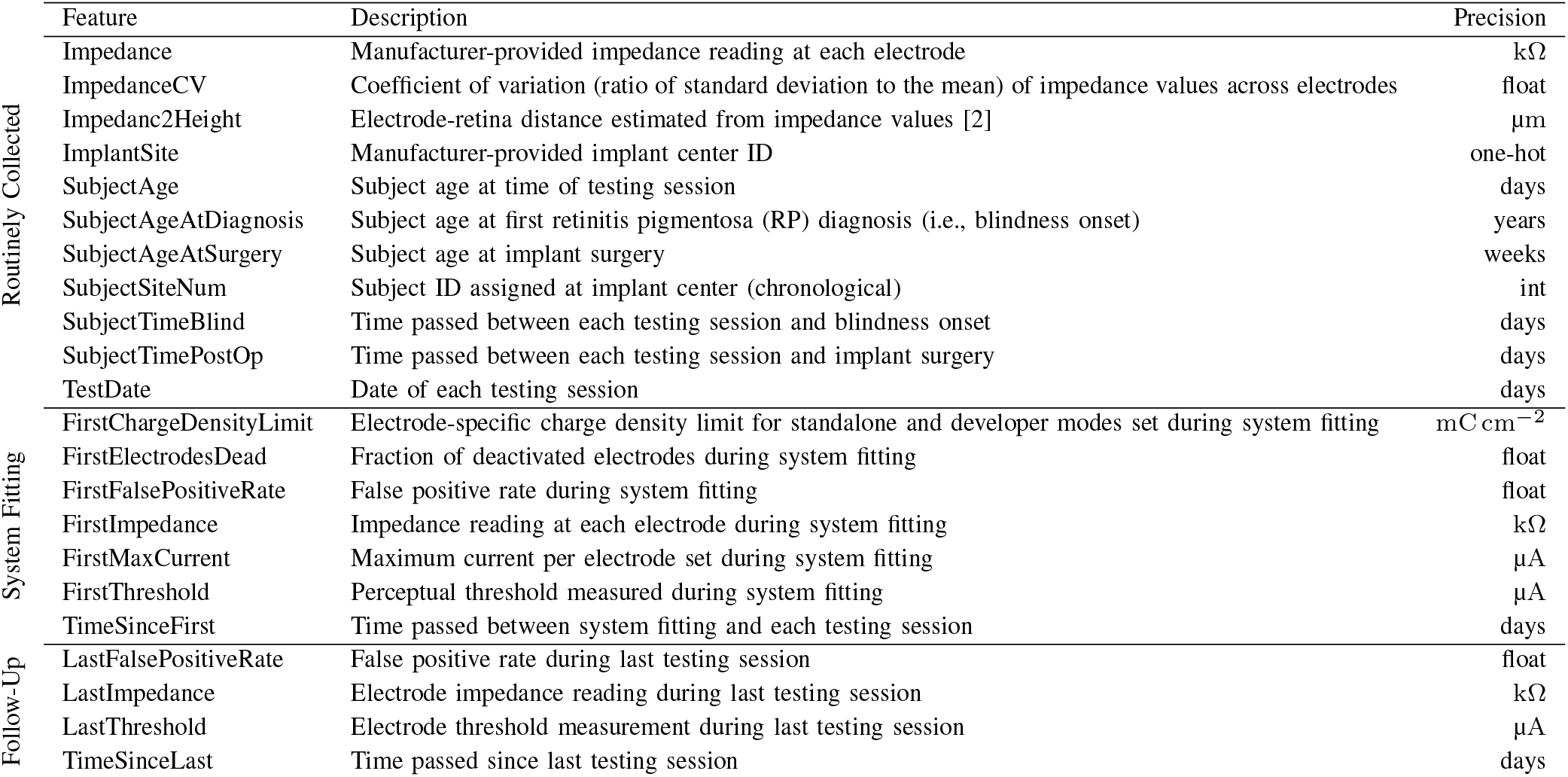
Feature Descriptions

As some parameters are more easily collected than others, a major goal of this work is to identify which of the available parameters are worth collecting for the purpose of predicting electrode deactivation. We therefore split the available features into three different categories:

1. *Routinely Collected Data:* We crowd-sourced public information about patient history (e.g., age at blindness diagnosis, age at implant surgery) from previous studies [3], [10], [11]. Surgery dates were obtained from Second Sight, and where not known exactly, were triangulated from the known dates of the earliest available impedance measurements and the 3-month follow-up exam. Knowing surgery dates and the subject’s age at that time allowed us to estimate the birth year for each subject (*±*1 year). Based on the above information and the dates of each testing session we were therefore able to calculate for each testing session: i) time since surgery, ii) time since diagnosis, and iii) subject age. Previous work identified electrode-retina distance as a key factor affecting thresholds [2]–[4]. Unfortunately, we did not have access to optical coherence tomography (OCT) images for all subjects. Instead we followed de Balthasar *et al*. [2] to estimate electrode-retina distance from the available impedance measurements (‘Impedance2Height’ in Table II).
2. *System Fitting:* Soon after system activation, patients undergo a system fitting procedure during which electrode impedances and perceptual thresholds are measured on all 60 electrodes. These values are then used to set several system parameters, such as the charge density limit and the largest allowable current. By default, charge density limits are set to 0.35 mC cm^*−*2^ per phase for everyday use, and to 1 mC cm^*−*2^ for lab use, but can be reduced for electrodes that are particularly sensitive. Analogously, stimulating currents are limited to 1 mA per default, but can be reduced for sensitive electrodes. Electrodes whose impedance value indicate either a short or open circuit are immediately deactivated. Thresholds and impedances obtained during system fitting were only used as features for the training data, not as labels that the algorithm was supposed to predict. We engineered several additional features, such as the fraction of deactivated electrodes, the coefficient of variation for impedance values measured across all electrodes, and the false positive rate during threshold measurements (where the stimulating current was zero but the patient reported seeing a phosphene). Finally, for each data point in the dataset, we calculated the time that had passed since system fitting.
3. *Follow-Up Examinations:* Patients participating in the Argus II Feasibility Protocol regularly visited their eye clinic for follow-up exams. We wondered how useful these more recent threshold and impedance measurements were for predicting electrode deactivation. Since the time between sessions varied, we also calculated the time that had passed since the last examination (‘TimeSinceLast’ in Table II).

### C. Explainable Machine Learning Model

We used gradient boosting (XGBoost), a powerful ML model based on an ensemble of decision trees [12], to predict electrode deactivation as a function of the model parameters (‘features’) described above. In this model, a strong predictor is built from iteratively combining weaker models (i.e., shallow decision trees). XGBoost has achieved state-of-the-art results in a variety of practical tasks with heterogeneous features and complex dependencies. To apply XGBoost to our longitudinal data, we assumed that each testing session was independent and treated timestamp-related data (e.g., ‘TestDate’, ‘SubjectTimePostOp’) as additional feature attributes.

To determine the relative importance of the different features for each model prediction, we adopted SHapley Additive ex-Planations (SHAP) [13]. SHAP is a feature attribution technique based on the game-theoretically optimal Shapley values, which determine how to fairly distribute a ‘payout’ (i.e., the prediction) among model parameters. SHAP first calculates a (local) feature importance value for each feature in a decision tree, and then accumulates these values across all trees in the XGBoost model to yield a global importance value for each feature.

Model performance was compared against two baselines: logistic regression (LR) with an L2 penalty and a support vector machine (SVM) with a linear kernel. Both baseline models were implemented using scikit-learn (v0.22.2.post1) [14] and Python 3.6.9.

### D. Model Evaluation and Comparison

To allow for a fair comparison between models, we used a nested leave-one-subject-out cross-validation, where we repeatedly fit the model to the data from all but one subject (outer loop). This procedure is equivalent to calculating the Akaike Information Criterion that takes into account the difference in number of parameters across models [15]. To tune the various hyperparameters of each model (LR: `C`; SVM: `gamma`; XGBoost: e.g., `max_depth`, `min_child_weight`), we split the training data again using leave-one-subject-out cross-validation (inner loop).

Model performance was evaluated on five common statistical indicators: accuracy (number of correct predictions), precision (number of correctly predicted electrode deactivations divided by the number of all deactivations), recall (number of correctly predicted deactivations divided by the number of all electrodes that should have been identified as deactivated), F1 score (harmonic mean of the precision and recall), and the area under the ROC curve (AUC).

## III. RESULTS

The cross-validated classification results are shown in Table III. Each model was trained on three different feature splits of the data as described in Table II: 1) relying solely on routine clinical measures (labeled ‘Routine’), 2) relying on routine measures and system fitting (‘Fitting’), and 3) relying on routine measures, system fitting, and data from follow-up examinations (‘Follow-up’).

**TABLE III.** Cross-Validated Classification Results

|  | Method | Accuracy | Precision | Recall | F1 | AUC |
| --- | --- | --- | --- | --- | --- | --- |
| Routine | XGBoost | **0.608** | **0.334** | 0.547 | **0.415** | **0.547** |
|  | SVM | 0.348 | 0.249 | **0.776** | 0.377 | 0.497 |
|  | LR | 0.483 | 0.264 | 0.579 | 0.363 | 0.485 |
| Fitting | XGBoost | **0.753** | **0.507** | **0.932** | **0.657** | **0.840** |
|  | SVM | 0.638 | 0.380 | 0.677 | 0.488 | 0.705 |
|  | LR | 0.681 | 0.430 | 0.782 | 0.555 | 0.799 |
| Follow-up | XGBoost | **0.840** | **0.634** | **0.873** | **0.734** | **0.903** |
|  | SVM | 0.830 | 0.619 | 0.853 | 0.718 | 0.897 |
|  | LR | 0.826 | 0.614 | 0.846 | 0.711 | 0.897 |

SVM: support vector machine, LR: logistic regression

Gradient boosting outperformed the two baseline models in all cases. Interestingly, without ever measuring perceptual thresholds (‘Routine’), XGBoost was able to predict future electrode deactivations with 60.8 % accuracy. In this case, the model’s decisions relied on initial impedance measurements as well as clinical information about the subject’s age, time since blindness onset, and time since device implantation. By incorporating threshold measurements as well as other parameters typically collected during system fitting, performance increased to 75.3 %. When additional measurements from follow-up examinations were included, XGBoost reached its peak performance (84 % accuracy, 0.903 AUC).

The SHAP values for the ten most important features in the dataset are shown in Fig. 2. In this plot, each data point is a prediction of electrode deactivation from the held-out cross-validation fold (test set). SHAP values indicate each feature’s contribution to the model’s decision, with positive values indicating that a feature pushed the model towards predicting deactivation, and negative values pushing the model away from predicting deactivation.

**Fig. 2.**
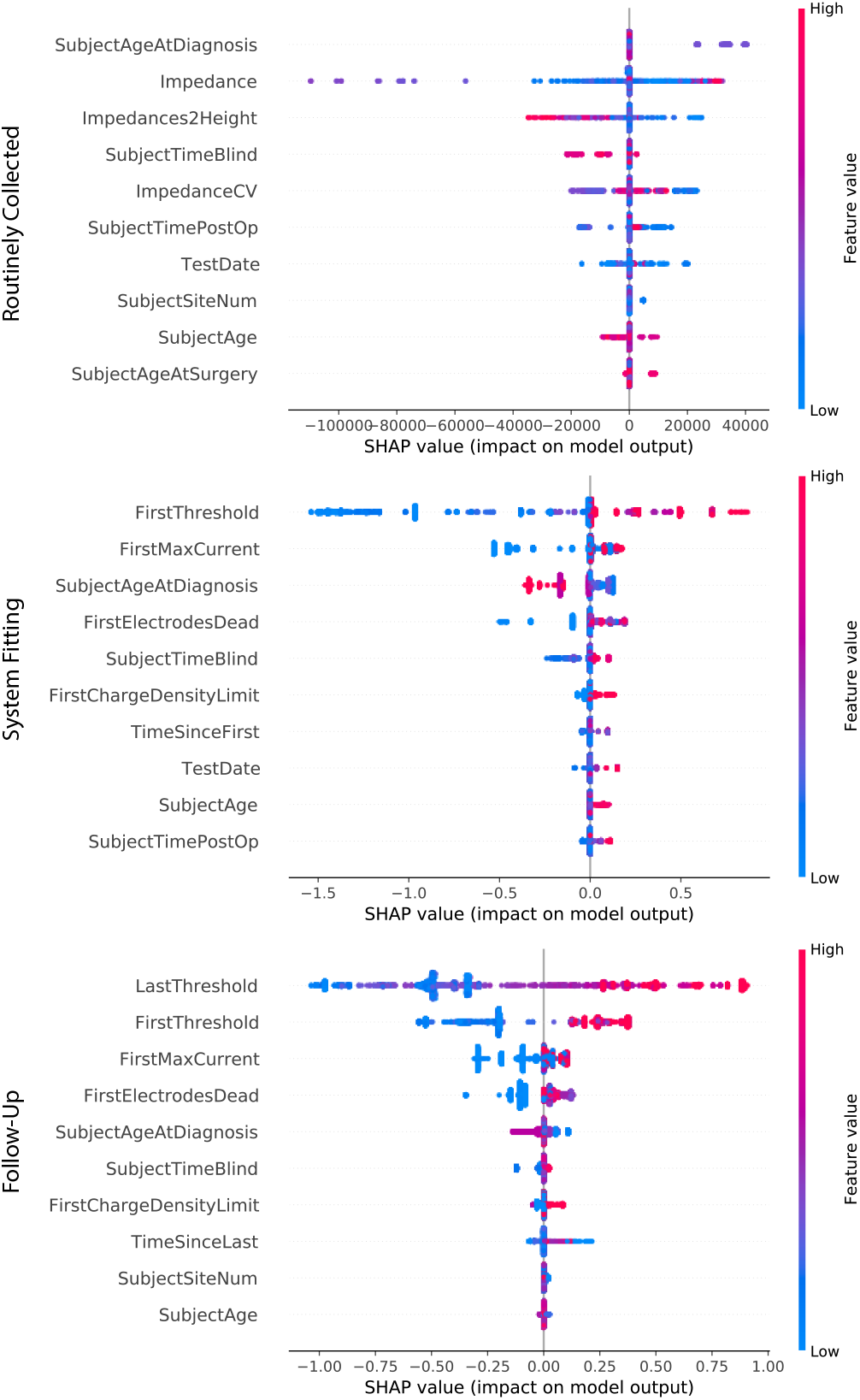
Force plot of SHapley Additive ex-Planations (SHAP) values for the ten most important features in the dataset (sorted top to bottom). Each data point is a prediction of electrode deactivation from the held-out cross-validation fold (test set). SHAP values indicate each feature’s contribution to the model’s decision (positive: pushing the model towards predicting deactivation; negative: pushing the model away from predicting deactivation). Colors indicate feature values. Results are shown for three different feature splits of the data (see Table II) including routinely collected measures, data from system fitting, and data from follow-up examinations.

Of all the routine clinical measures (top panel in Fig. 2), subject age at diagnosis, impedance, and an estimate of retinal-electrode distance proved the most important. Other influential factors included time since blindness onset, time since device implantation, and subject age.

Threshold measurements and related electrode-specific settings (e.g., maximum current, charge density limit) typically obtained during system fitting proved to be even more important (middle panel in Fig. 2). The higher the initial threshold, the more likely an electrode was to be deactivated in the future. The model also identified the overall number of electrodes deactivated during system fitting as an important predictor of future deactivations.

Not surprisingly, when data from follow-up examinations was considered as well (bottom panel in Fig. 2), the most recently obtained threshold measurement proved to be the most important feature. This is consistent with the finding that threshold measurements often go through large fluctuation over time [5], which cannot be predicted from an initial threshold measurement during system fitting.

In all three scenarios, parameters such as subject age and time since blindness onset proved to be highly predictive of future electrode deactivations.

## IV. DISCUSSION

We found that an XAI model could predict electrode deactivation from routine clinical measures with 60.8% accuracy. Performance increased to 75.3% when system fitting data was available, and to 84% when thresholds from follow-up examinations were available. On the one hand, these findings highlight the importance of periodical threshold measurements to continuously monitor device performance. On the other hand, in the absence of such measurements, our work demonstrates that routinely collected clinical measures and a single session of system fitting might be sufficient to inform an XAI-based electrode deactivation strategy.

Unfortunately, we did not have access to OCT and fundus images, which would have allowed us to infer anatomical parameters such as electrode-retina distance and retinal thickness. However, even if available these images are often of limited use [3]: most retinal implant recipients present with nystagmus, and electrodes cast shadows on the OCT b-scan. This complicates the readout of parameters such as retinal thickness and electrode-retina distance.

Consistent with previous studies [3], [4], we found that electrode impedance is an important predictor of perceptual thresholds, and thus electrode deactivation. The model further identified subject age and time since blindness onset as important predictors of future electrode deactivations. As advanced cases of retinitis pigmentosa (RP) are associated with worsening visual outcomes, it is perhaps not surprising that time since blindness onset could serve as a proxy for disease progression. These results suggest that parameters such as age and time since blindness onset may be important for predicting visual outcomes with patient-specific computational models of prosthetic vision [16], [17]. However, as our study is limited to Argus II data, future work should focus on replicating these results based on data from other (and preferably: multiple) retinal implants.

To the best of our knowledge, this is the first systematic study of electrode deactivation for the field of retinal prostheses. An accurate predictive model of electrode deactivation that relies on routine clinical measures has the potential to benefit both the retinal implant and wider neuroprosthetics communities. In the near future, such data-driven approaches could complement expert knowledge–driven interventions [18], making increasingly few assumptions about the underlying data and instead automatically inferring and exploiting relationships among the measured features.

## Data Availability

N/A

## Notes

### Competing Interest Statement

The authors have declared no competing interest.

### Author Declarations

Our study, which did not involve human subjects research, was exempt from IRB approval.

